# Spectral Analysis of the T Cell Repertoire Recovery Following Allogeneic Stem Cell Transplantation

**DOI:** 10.64898/2026.09.28.26364185

**Authors:** Amir A. Toor, Alejandro Marinos-Velarde, Rehan Qayyum

## Abstract

The human T cell repertoire is generated through recombination of a multitude of T cell receptor gene segments, yielding a complex array of T cell clones. T cell receptor beta (TRB) V gene segment defined, T cell clonal frequency when organized in correspondence with the respective V gene segment positions on the TRB genomic locus yields a periodic, undulating curve in the spatial domain. Using the genomic distance from the TRB-D1 segment to the TRB-V1-29 segments, spectral analysis was performed utilizing Lomb-Scargle periodogram (modified Fourier analysis) to obtain spectral power curves quantifying the TRB V clonal frequencies from 12 allogeneic stem cell transplant donors (baseline) and recipients (<100 days or b/w 100-365) using a variety of analytic software. Patients either underwent HLA matched related bone marrow or HLA matched unrelated blood stem cell transplantation, using PT-CY for the former and ATG for the latter. Spectral Power curves revealed dominant spectral peaks at wavelengths ranging from 113-335 millicycles/kb in the 12 donors, with consistent frequency domain spectral patterns, supporting similar use of V segments across healthy individuals. PT-CY recipients had power spectra closely corresponding to the respective donors, however the ATG recipients had relatively dispersed spectra, with spectral centroid shifted towards higher frequencies compared to donors and a reciprocal decline in the low frequency indices. Spectral power was concentrated in the <3 kb and 3-12 kb wavelengths in both groups, consistent with the periodicity observed in the relative V gene segment usage across the repertoire. The analyses reported here demonstrate that the healthy SCT donors have a spectral signature occupying short to intermediate wavelegnths in the frequency domain, PT CY recipients recover with donor-like spectra on the average, however ATG recipients tend to shift towards higher frequencies. These findings are consistent with a normal organized distribution of TRB V segment usage in healthy individuals, with PT-CY preserving this organization more consistently than ATG. Spectral (Fourier) analysis of TRB sequencing data provides a repertoire wide summary of T cell clonal distribution.

## Introduction

Contemporary allogeneic stem cell transplantation has been revolutionized by the administration of post-transplant cyclophosphamide (PT-CY) to patients undergoing transplantation across the continuum of HLA match grades. The risk of graft versus host disease and the need for prolonged immune suppression are diminished, without a commensurate increase in the risk of relapse when patients with hematological malignancies are transplanted. There is a risk for infections, nevertheless across multiple studies involving both HLA matched and mismatched donors the risk for graft versus host disease (GVHD) – relapse free survival (GRFS). The mechanism for this modulation of alloreactivity is attributed to elimination of alloreactive T cells and the emergence of regulatory T cells which enhance the tolerogenic potential of the allografts. Other means of donor T cell depletion such as rabbit anti-thymocyte globulin are effective at diminishing chronic GVHD without impacting relapse risk. Finally, traditional GVHD prophylaxis approaches are fraught with too high a chronic GVHD risk to be of broadly feasible except in the case of best HLA-matched donor-recipient pairs (DRP). Considered in aggregate, the GRFS observed in clinical trials studying PT-CY remains consistently higher than other GVHD prophylaxis modalities across the spectrum of DRP. It is likely that this phenomenon may be the result of restoration of a normal, healthy T cell repertoire following transplant and the lower intensity and shorter course of post-transplant immune suppression in the majority of PT CY treated patients. Comprehensive analysis of T cell repertoire recovery is therefore a critical rea of study.

T cell repertoire recovery is best measured by quantifying the T cell clones present in circulation at any given time following SCT. This is accomplished by sequencing of the T cell receptors (TCR) alpha and beta using various next generation sequencing (NGS) platforms. These yield immense data sets, with millions of TCR-alpha/beta sequences, along with their frequency. The complexity of these data sets limits their utility in day to day clinical practice, making TCR sequencing primarily a research tool. These data are summarized using analytic methodology from information theory, utilizing such concepts as T cell richness/evenness and Shannon’s entropy to represent large data sets in a compact fashion and subject TCR clonal frequency data from different patients to statistical analysis and to draw appropriate inferences regarding the clinical impact of T cell repertoire recovery. Nevertheless, by analyzing individual clonal frequencies as a function of the entire T cell population, these indices yield summary values which do not truly reflect the underlying biological complexity. This results in loss of information, rendering the ensuing correlations approximate at best. Therefore, alternative analytic methodology is needed to understand TCR repertoire data, which may more completely reflect the underlying data structure and provide an intuitive, quantitative understanding of T cell repertoire evolution over time in different clinical scenarios.

The human T cell repertoire has several organizational principles which have been previously elucidated and may be utilized in developing new methods for studying repertoire diversity. First, TCR defined clonal frequencies are logarithmically scaled and have Power Law distributions. Inherent to this quality, when clonal frequencies are assessed from a TCR-beta (TRB), Variable, Joining and Diversity (VDJ) segment defined clonal frequency perspective, there is a fractal ordering present, i.e., distribution of T cell clones remains proportional across different levels of V,D and J segment recombination.^1^ Second, when the T cell receptor V, D and J segment locations are examined on the TCR loci, these are highly ordered, with segment lengths and the inter-segment distances (in base pairs) also being logarithmically scaled. V gene segments are the longest, J gene segments have an intermediate length and D gene segments are the shortest, with corresponding, proportional inter-segmental distances. Third, when the TCR beta defined clonal frequencies are examined as a function of the V and J segment positions on the TCR beta locus on chromosome 7q,^2^ a periodic distribution of clonal frequencies is observed as the clonal frequencies are mapped across the TCR beta locus from the 5’ to the 3’ end of the locus.^3, 4^ Last, minor histocompatibility antigen – HLA binding affinity distributions mirror T cell clonal distribution, enabling the generation of mathematical models, based on the vector operator systems which allow consideration of human immune responses as dynamical systems. ^5, 6^ Given the mathematical organization of the repertoire and that immune responses following stem cell transplantation (SCT) behave as dynamical systems,^7, 8^ logically the entire T cell repertoire should behave in a broadly quantifiable manner across the entire T cell receptor array. Such broad quantitative characterization of the T cell repertoire may permit a deeper understanding of large-scale phenomenon observed post-transplant.

Fourier analysis is a method of analyzing periodic phenomenon, such as waves travelling across different media (e.g., soundwaves propagating through air). The underlying assumption is that complex waveforms (such as a multi-instrument melody) represent a sum of many simple sine waves (fundamental harmonics) which combine to yield the final note. In other words, several waves of different frequency (number of oscillations over unit time) of a certain amplitude (magnitude) combine to yield a complex wave. ECG or EEG patterns are similar periodic phenomenon with complex waveforms. Fourier analysis is a mathematical method which allows the complex waves to be deconstructed into constituent simple harmonic waves of different frequencies, and calculates the contribution of each unique frequency to the overall amplitude of the combined wave. This is accomplished by a mathematical procedure called Discrete Fourier Transformation (DFT). The DFT takes the amplitude (height) of a wave from the time domain (amplitude plotted on the y axis, as a function of time on the x axis), and calculates the contribution of each constituent frequency comprising the complex wave: in turn plotting the amplitude in the frequency-domain (the magnitude contribution of each frequency to the final amplitude of the complex wave, Power). The relevant frequencies comprise the spectrum encompassed by the complex waves. As an example (**Figure**) two simple waves (Blue: low frequency/long wavelength, Red: High frequency/short wavelength) combine to form a complex wave (Purple combined bimodal wave), which when transformed and visualized in the frequency domain can be seen to be comprised of two waves of different frequencies (thus different wavelengths) contributing different amplitudes to the final wave. This comprises the frequency spectral representation of the complex wave.

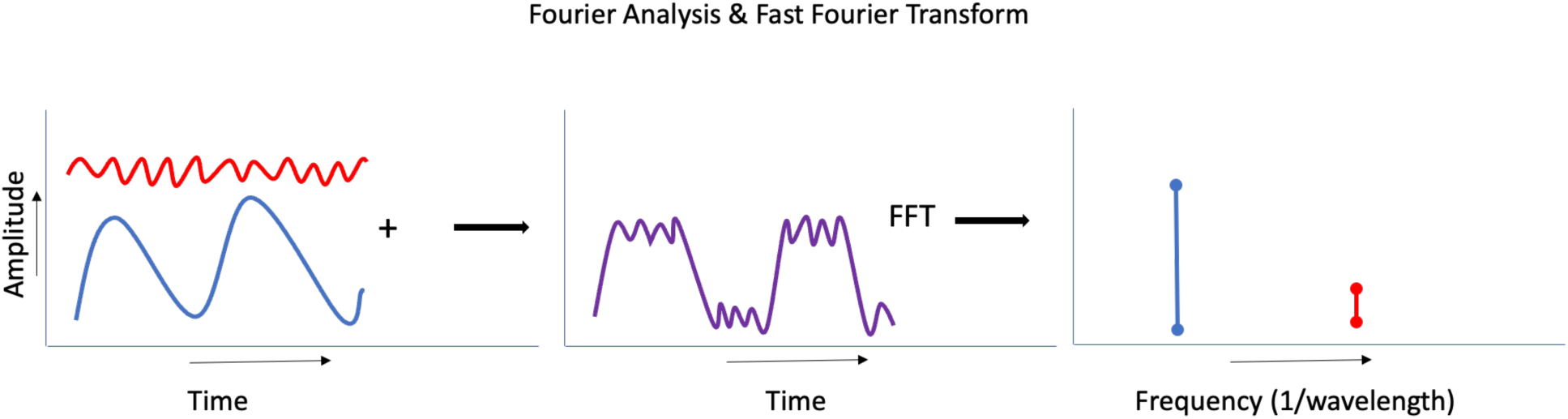

This concept of oscillation across the time domain to produce a periodic phenomenon (wave) may be similarly extrapolated to any cyclic variation in phenomenon with a measurable output, such as the probability of VDJ recombination occurring across a strand of double helical DNA molecules with V and J genes sequentially organized on it; in this instance the periodic phenomenon of interest is variation in the probability of TRB V and J genes participating in recombination and contributing to the final repertoire (represented by T cell clonal frequencies), producing oscillations mapped to a spatial domain, such as the TRB locus (measured in base-pairs of DNA). Like time domain-based phenomena (sound waves), this spatial waveform quantifying the sequential V gene segment defined T cell clones, may also be examined in the frequency domain to estimate the relative contribution of V gene segments across the TRB locus to the final T cell clonal repertoire. Therefore, when the T cell repertoire at different time points is measured, any change in the relative TRB (or TRA) locus distribution of gene segment usage may be summarized and information regarding T cell clonal evolution be summarized, with relatively greater preservation of information than single, numeric measures. Fourier analysis does so by decomposing the final T cell repertoire to highlight the relative contribution of different regions of the TRB gene. Applied to large data sets, this method may prove to be a versatile tool for comparing T cell repertoire both within and between individuals and over time. In this paper, an adjusted Fourier analysis of TRB sequencing data from two distinct patient cohorts is presented as proof of principle.

## Methods

Archival, anonymized data from patients who underwent allogeneic peripheral blood stem cell transplantation on IRB approved protocols from HLA matched donors at the Virginia Commonwealth University (VCU) and John Hopkins University (JHU) were utilized.^9, 10, 11^ SCT donor and recipient samples for determining T-cell clonal frequency were obtained as part of a clinical trial approved by the institutional review boards at VCU (ClinicalTrials.gov Identifier: NCT00709592) and JHU (ClinicalTrials.gov Identifier: NCT00809276). The VCU cohort underwent HLA matched unrelated donor transplantation with GCSF mobilized blood stem cells and GVHD prophylaxis utilizing rabbit ATG, tacrolimus and MMF (henceforth ATG cohort); the JHU cohort underwent HLA matched related donor bone marrow transplantation utilizing PT CY, CsA and MMF as GVHD prophylaxis (henceforth PT-CY cohort). In the ATG cohort T cell repertoire evaluation was performed on complementary DNA isolated from the donor product, and the recipient blood samples at ≥ day 100 upto one-year post transplant, using Adaptive Biotechnologies’ T cell receptor Clonoseq assay (Adaptive Biotechnologies, Seattle, WA).^1, 3^ In the PT-CY cohort the TRB sequencing was performed utilizing genomic DNA isolated from donors, and recipients samples at <100 days (Early) and ∼365 days (Late).

All the specific TRB-V gene sequence reads in each individual were summed and tabulated sequentially according to the 5’ to 3’ order of the V gene segments on the TCR beta (TRB) locus, along with the genomic distance of the respective loci from the TRB D1 gene segment. A frequency of zero was given to the TRB V pseudo genes. The spatial dimension of the TRB V gene segments was the angular distance (in radians) from the TRB D1 segment;^3^ approximated by KB for ease of understanding. Of note, TRB V30 is located 3’ of the TRB D gene segment so was excluded from the reported analyses, but included in sensitivity anlaysis. The ATG patients presented here underwent HLA MUD SCT (8/8, n=5; 7/8, n=1 – DRP 14/HLA B mm). Samples 3 and 5 were collected at day 100 (R2) and the remaining samples between days 100 - 365 (R3).

### Spectral Analysis Methodology

Spectral analysis was performed using a custom Stata pipeline (StataNow 19, StataCorp, College Station, TX) supplemented with Python 3 (scipy.signal.lombscargle, numpy, pandas, matplotlib) for numerical computation and visualization. The analysis was adjusted to apply the Lomb-Scargle periodogram approach to the unevenly spaced V-gene clonal frequency data, avoiding the interpolation artifacts that can arise with conventional Fast Fourier Transform (FFT) methods.

For each sample (30 total: 12 donors (ATG and PT CY n=6 each; 18 recipients ATG n=6, PT-Cyn=6 ear;y and 6 late), the 65 TRB V gene segment defined clonal frequencies (copy number) were arrayed according to the genomic position of the respective gene segments on chromosome 7q, measured in radians (calculated form kilobases (kb) of DNA base pairs) from the TRB D1 gene segment. For pseudogenes a value of 0 was assigned for the clonal frequencies. The ATG cohort also included TRB-A and TRB-B pseudogene segments in the initial analysis, these were removed for later pooled analysis. The genomic coordinates of the V gene segments are inherently unevenly spaced, reflecting the physical organization of the TRB locus. The Lomb-Scargle periodogram computes spectral power at each candidate frequency by performing a least-squares fit of a sinusoidal model to the raw data, making it suitable for unevenly sampled spatial data without requiring interpolation. The spectral power sums to 1 across all the frequencies derived for each individual repertoire examined.

The frequency grid was defined from a minimum frequency f_min_ = 1/60 kb (0.016 cycles/kb [16 millicycles/kb]; corresponding to a maximum wavelength of 60 kb) to a maximum frequency f_max_ = 0.5 cycles/kb [500 millicycles/kb] (0.5 cycles/kb; wavelength 2 kb; Nyquist limit for the minimum inter-V-gene spacing). The frequency grid consisted of 1024 evenly spaced bins on the linear frequency scale. The relevant section of the TRB locus between TRB-V1 and TRB-D1 spans ∼550 kb. The standardized frequency window enables reproducibility across analyses by fixing the long-wavelength cutoff at 60 kb, rather than using a data-dependent cutoff such as 1/(genomic span), which varies with the number of V genes included. An important consideration here is that unlike time dependent periodic phenomenon, the TRB clonal frequency waveform has no negative component, with all numeric values going from 0 to *x*. It is also important to recognize that the frequency spectrum represents a compilation of calculated spatial frequency bands which summed together, yield the TRB V gene segment defined T cell clonal frequency distribution across the locus. Therefore these bands taken together represent the relative contribution of the V gene segments across than the entire TCR locus.

### Spectral Metrics

The following metrics were derived from each power spectrum. The spectral centroid (SC) is the weighted mean frequency of the power spectrum, calculated as Sum (f_i * P_i) / Sum(P_i), where f_i is the frequency and P_i is the power (amplitude, or T cell clonal copy numbers) at that frequency. In essence the Fourier analysis determine the frequency spectrum which best represents the repertoire; SC captures the central tendency of the spectral distribution. Higher SC values indicate power concentrated at shorter wavelengths (higher frequencies). This would imply relative flattening of the TRB repertoire periodicity, or a diminution of the periodicity observed, with more uniform contribution of TRB V gene segments. The low-frequency index (LFI) is the fraction of total spectral power below 100 millicycles/kb (10 kb wavelength). A higher value will be consistent with a more restricted, albeit periodic TRB gene segment use and reflect relative oligoclonality. Spectral entropy is the Shannon entropy of the normalized power distribution, measuring the uniformity of power spread across the frequency grid, with higher values corresponding with greater randomness across the frequency spectrum, corresponding to a broadly distributed repertoire in terms of V gene segment contributions. Band power is the fraction of total repertoire contribution (spectral power) within defined wavelength intervals: 3-12 kb (Short Spacing), 12-30 kb (Intermediate Spacing), 30-60 kb (Long spacing), and (Ultra-long pacing) >60 kb. The residual band (<3 kb) captures power at the highest frequencies. In essence this indicates the calculated frequency bands which contribute to the greatest extent in the final T cell repertoire generation.

For the ATG cohort, the analysis was repeated using MATLAB R2026a (The MathWorks, 2026) with the Signal Processing Toolbox add-on. Spectral analysis was performed using the Lomb-Scargle algorithm to identify periodicities in the data. Fast Fourier Transform was not used since genes are not evenly spaced in the genome. Signal intensities for both patient and donor datasets were standardized using z-score normalization (mean = 0, standard deviation = 1) prior to analysis to control for amplitude differences between samples. The reported parameters were spectral centroid, and Shannon’s entropy.

The complete analysis pipeline is implemented in the do file fft_dr_reproduction.do, which is available as supplementary material. The full Stata log and all output tables are provided in the supplement.

## Results

### Periodicity in TRB V defined clonal frequency

The TRB V clonal frequency distribution when organized across the TRB-V locus (V1-V29) demonstrated periodicity in the spatial domain for all samples as previously reported. It was evident that the TRB-V defined clonal frequency from the patients in the ATG cohort had significant variation between donors and recipients but occupied the same scale (**Figure 1**). On the other hand, TRB V clonal frequency in the PT-CY patients maintained the same pattern, only diminishing in scale by orders of magnitude in the recipients.

**Figure 1.**
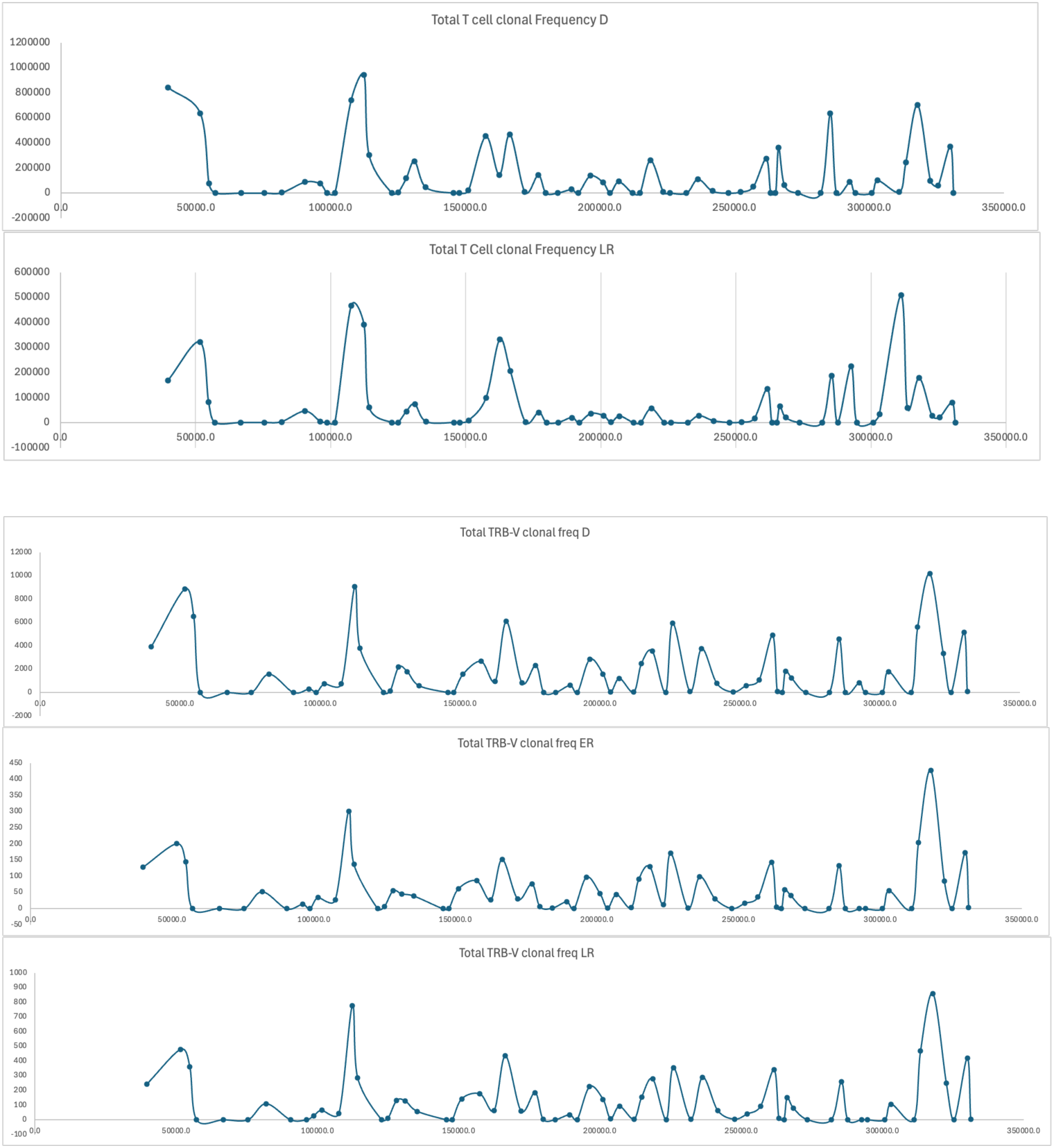
Periodic distribution of the total TRB V gene segment defined clonal frequency across the TRB V gene segments in the spatial domain. Top two panels, representative data for ATG cohort DRP (DR9), bottom 3 panels for PT-CY (001-023). D-donor, ER and LR-early & late recipient. X-axis, Angular distance from TRB-V1 to TRB-D1 in radians (bp_n_*2π/10) and Y-axis, total number of reads, note scale variation in the different graphs.

### Spectral analysis of TRB V defined cell clonal frequency

To obtain the frequency domain representation for these data, Lomb-Scargle periodogram analysis was performed on TRB V gene clonal frequency data from the ATG and PT-CY cohorts (n=6 DRP each), yielding 30 power spectra sampled on a 1024-bin frequency grid with a standardized frequency window (f_min_ = 1/60 cycles/kb, f_max_ = 0.5 cycles/kb). Decomposition of this spatial waveform into its constituent frequencies via the Lomb-Scargle periodogram revealed dominant spectral peaks at wavelengths of approximately 113-252 millicycles/kb for the donors in the ATG cohort vs. 115–335 mcycles/kb for the PT CY cohort, corresponding to the spacing between V gene clusters on the TRB locus. The most striking feature was the homogeneity observed in the spectra of donors, consistent with a similar usage of TRB V gene segments across the normal donors studied, despite the different donor characteristics in the two cohorts (**Figure 2 & Figure P1**). The recipient spectra for the ATG cohort demonstrated greater heterogeneity in their frequency distribution, in contrast to the PT CY spectra which similar to the donor spectra for both the early (<100 days post-transplant) and the late (∼1 year) time points (**Figure 3 & Figure P2**).

**Figure 2.**
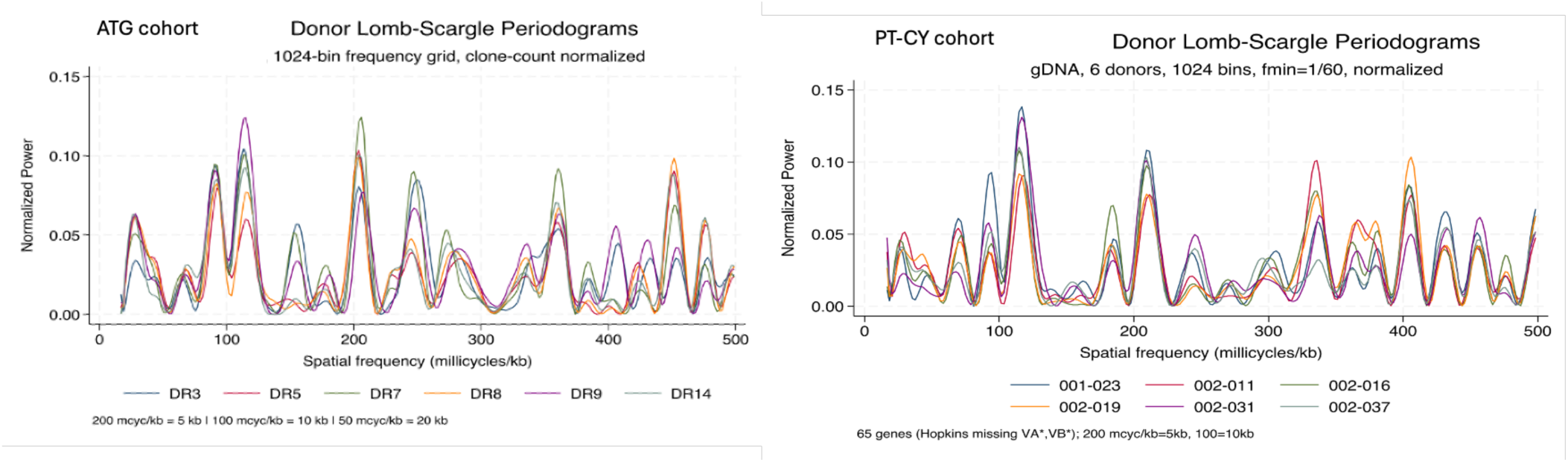
Donor Lomb-Scargle power spectra for all donor-recipient pairs in the two cohorts. The frequency axis spans 16.7 to 500 millicycles/kb. Power spectra are normalized to total power = 1. The 6 Donors show a conserved spectral signature with dominant power in the 110-250 millicycles/kb range (corresponding to 4-9 kb wavelength).

**Figure 3.**
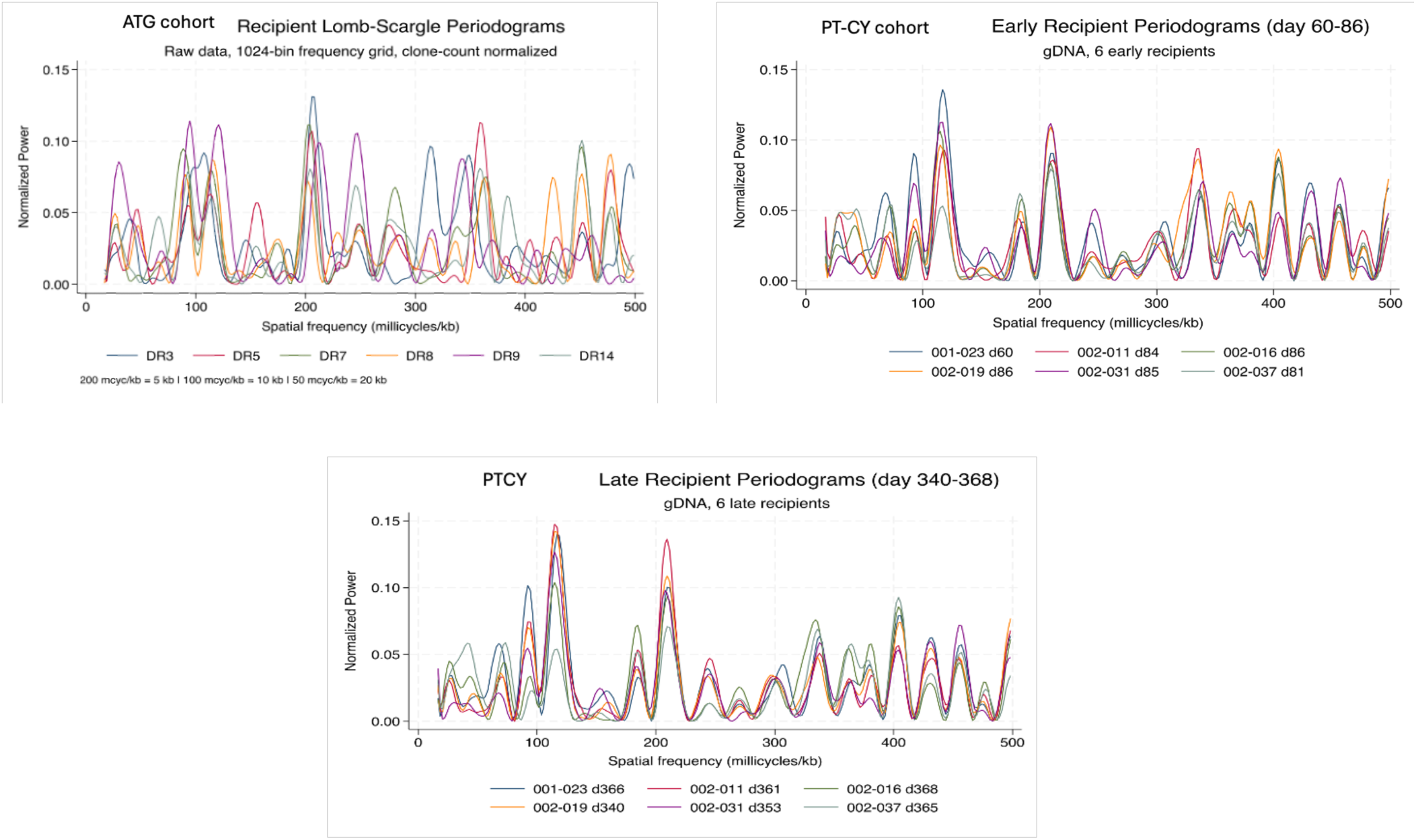
Overlay of the recipient power spectra. While Donor spectra (Figure 1) show tight clustering, indicating a conserved spectral signature across individuals, ATG recipient spectra show greater dispersion, reflecting variable post-transplant reconstitution while the PTCY recipient spectra approximated their donors at both time points. In the ATG cohort, recipients diverge from their matched donors to varying degrees, shifting towards longer wavelengths.

The magnitude of donor-recipient spectral power divergence in the two cohorts was different between ATG and PT CY recipients (**Figure 4 & Figure P3**), implying differential V segment usage in the recipient as the T cell repertoire reconstitutes following transplantation. Whereas ATG recipients diverged markedly from their respective donors across their frequency spectra, the variance was considerably dampened in the PT-CY cohort.

**Figure 4.**
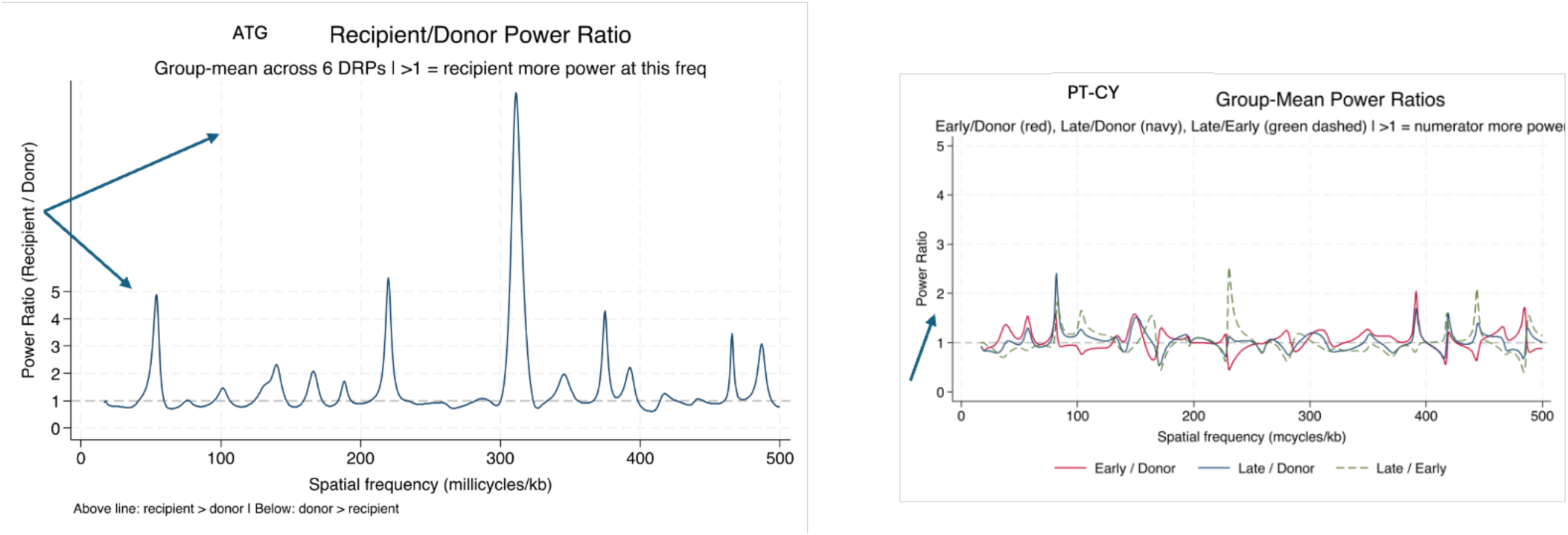
Donor-recipient power ratio across the frequency spectrum. Isolated group-mean power ratios, values >1 indicate relatively more power in the recipient at that frequency. In the ATG cohort, recipients carry approximately 2-3x more power than donors at very short wavelengths (3-5 kb) and at 20-40 kb. In the PT-CY cohort Early/Late vs. Donor and Late/Early oscillate around 1.0.

### Spectral Metrics

The primary spectral metrics computed for each sample are summarized in Table 1. In the ATG, URD BSCT recipients, the donor spectral centroid (SC) ranged from a relatively narrow range of 231.0 to 249.1 millicycles/kb (median 240), and a more dispersed, 213.6 to 264.9 millicycles/kb in the recipients (median 260), reflecting greater disorder and randomness in the usage of the V gene segments in the post-transplant reconstituting T cell repertoire. In 5 of 6 pairs, the recipient’s SC was higher than the paired donor’s (**Figure 5**), indicating a shift toward higher-frequency (shorter-wavelength) spectral components after transplantation. These findings support the notion, that in ATG recipients, the T cell repertoire has disruption in the normal periodic distribution observed in the donor T cell repertoire V gene representation in the T cell clones. In other words, the normal hierarchy of V segment usage is disrupted in the post-transplant setting. Correspondingly, the SC range in the PT-CY MRD, BMT recipients, the (SC) ranged from a relatively narrow range of 250.1 to 272.0 millicycles/kb in donors (median 258.4), and a relatively preserved, 249.6 to 267.7 millicycles/kb (median 260.4) in the early sample from the recipients, and 247.7 to 262.4 millicycles/kb (median 256.4) in the late sample. The preservation in the SC amongst the PT-CY, BMT recipients corresponds to the relative homogeneity observed across the power spectra of these DRP as opposed to the ATG, BSCT recipients.

**Figure 5.**
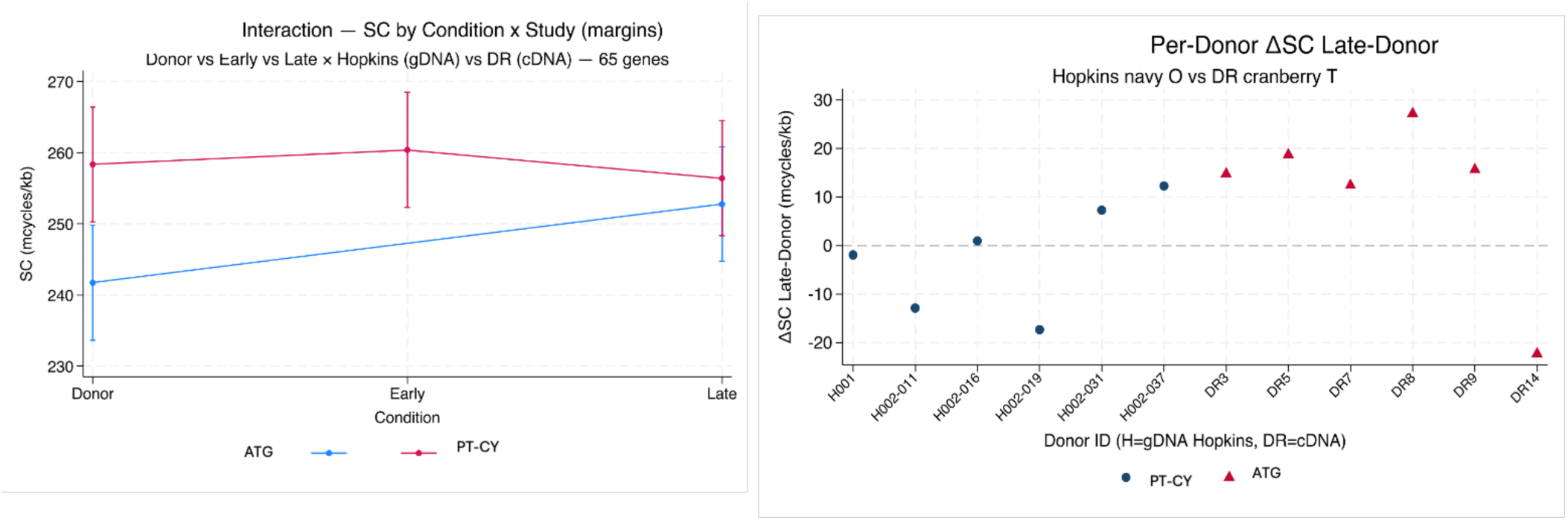
Spectral centroid comparison for each donor-recipient pair. PT CY (teal) vs ATG (navy) parallel lines: Donor→Late slopes similar after study offset, interaction NS, confirming additive study indicator adequate. Per-donor ΔSC Late-Donor dot plot (PT-CY navy O vs ATG cranberry).

Consistent with the higher frequency shift in the SC in the ATG cohort recipients (delta SC PT-CY −1.96 [−11.1,7.2] vs. delta SC ATG +11.06 [−2.65,24.76]), the decline in the low frequency index (LFI - fraction <100 mcycles/kb (λ>10 kb)) was reciprocally greater in the ATG recipients (delta LFI PT-CY −0.002, delta LFI ATG −0.025), and minimal in the PT-CY recipients.

Spectral entropy ranged from 9.41 to 9.54 bits in donors and 9.37 to 9.53 bits in the recipients in the ATG DRP, with no consistent donor-recipient direction (2 of 6 pairs showed increased entropy in recipients). For the PT-CY DRP the corresponding ranges were 9.44 to 9.53 bits for the donors, and recipients (early time point) and 9.40-9.49 bits for the late time points. The uniformly high entropy values across the cohorts (maximum possible for 1024 bins is approximately 10 bits) indicate that spectral power is broadly distributed across the frequency grid. Nevertheless, the ATG cohort had greater variability in recipient entropies, consistent with the more dispersed recipient frequency spectra observed in these patients (**Figure 6**). The broadly high entropy is likely a consequence of the normally complex structure of the T cell repertoire given the multiplicity of V genes involved under normal circumstances, and the afore mentioned complexity of the periodic V segment usage waveform.

**Figure 6.**
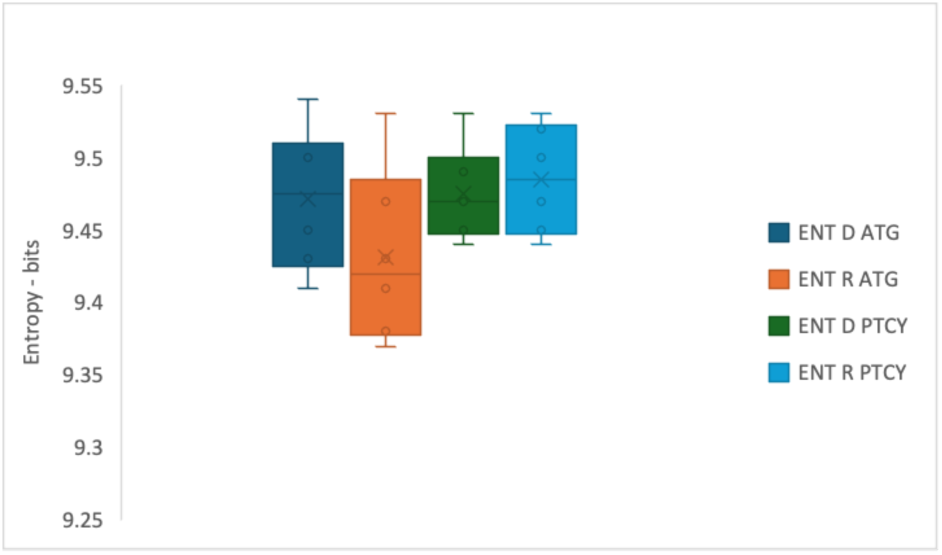
Spectral entropy in donors and recipients the two cohorts.

### Wavelength Spectra and Band Power Distribution

Distribution of power across biologically defined wavelength bands is an alternative representation of frequency spectra to highlight any differences in the repertoire structure across the cohorts. Amongst all twelve DRP, the 3-12 kb band (sub-family spacing) accounts for 55.9 +/−3.9% of total spectral power in the ATG/BSCT donors and 54.9+/−4.8% in recipients, while these values were 50.3+/−5.6% and 48.6+/−5.8% respectively in the early PT-CY/BMT cohort, going up to 52.1+/−8.7% in the late recipient samples (**Figure 7**). This suggests that the sub-family spacing pattern dominates the TRB V gene usage spectrum. The residual high-frequency band (<3 kb) accounts for 32.0 +/− 2.4% (donors) and 34.5 +/− 5.8% (recipients). There was no consistent aggregate difference in the band power representation across DRP in the two cohorts. The within-pair similarity of band power — despite individual differences in total clonal frequency and spectral centroid — suggests that the sub-family spacing pattern is a conserved property of the TRB repertoire architecture rather than a marker of repertoire perturbation. However, within each pair, within the bands, there emerged considerable heterogeneity in the ATG recipients, specifically, donor peaks split by platform, for the PT-Cy/BMT donors it was ∼8.5–8.7 kb (except for two donors, 002-011 at 3.0, 002-019 at 2.5 kb) vs ATG/BSCT donors it was ∼4.9–8.8 kb, both inside the 3–12 kb zone, which renders band power platform-robust compared with other spectral metrics, for example SC. In recipients of the ATG cohort, wavelength peaks shift to a shorter range (2.1–4.8 kb except an outlier, DR9 at 10.6 kb) while the PT-CY late sample cohort remains stable in the ∼8.5–8.7 kb (except, one, 002-011 converging 3.0→8.7 kb and another, 002-037 dropping to 2.5 kb). This dispersion of the dominant wavelengths observed in the ATG recipients suggests fine scale variation in the V gene segment utilization compared with the respective donors despite broader organizational similarity.

**Figure 7.**
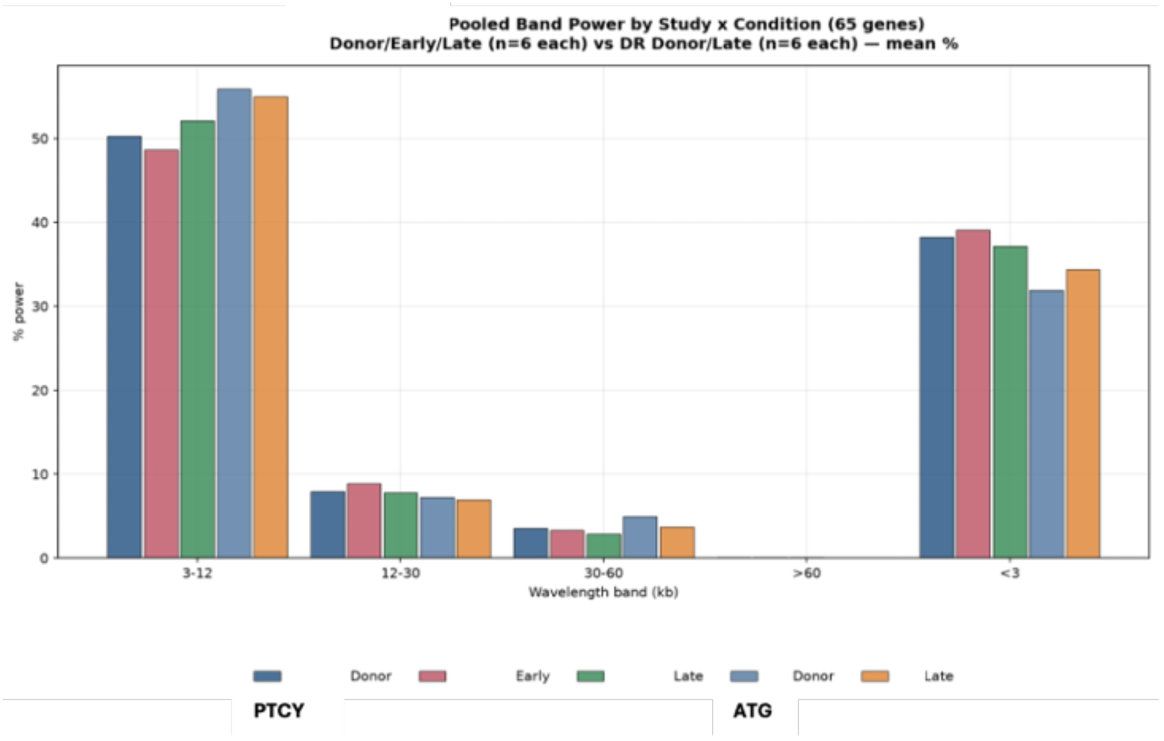
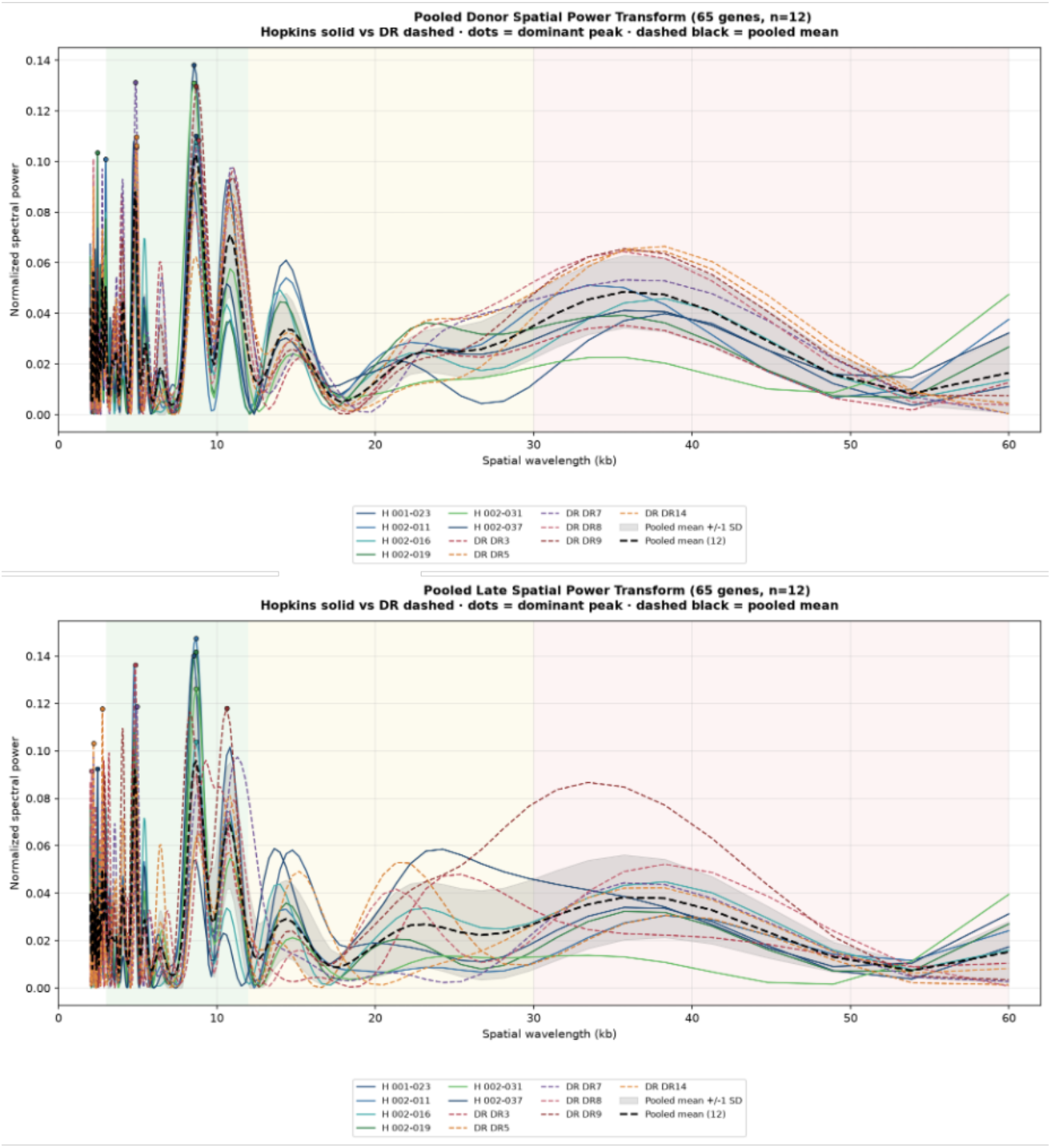
Spectral power distribution by wavelength band comparison between donors and recipients. Band distribution of the Power in dominant spectra. Dominant wavelength highlighted with closed circle. Wavelength Band and TCR V gene segment family’s assignment is as below.

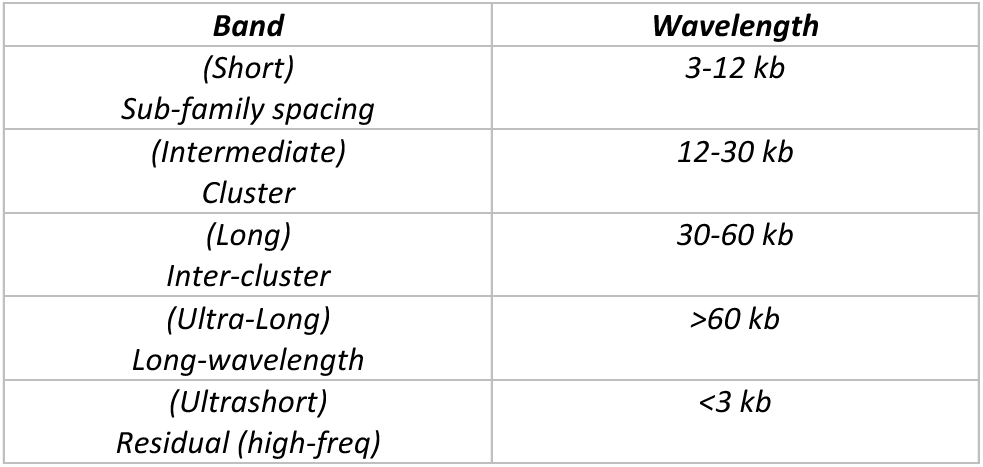

## Discussion

In patients with hematological malignancies, an allogeneic SCT is inherently dependent on the donor-recipient alloreactivity for its therapeutic graft vs. leukemia effect, however often this is complicated by graft vs. host disease. Immune suppression impedes T cell recovery, which in turn increases the risk for infection, as does the oligoclonal expansion of alloreactive T cells post-transplant. These effects impact donor-recipient pairs stochastically and are not predictable with a high degree of accuracy, given the heterogeneity inherent to the HLA system, and the human exome as a source of alloreactive minor histocompatibility antigens.^12^ T cells are the main protagonists for these immunological phenomenon and deep analysis of the T cell repertoire with NGS has been utilized to help understand immune recovery post-transplant and its influence on clinical outcomes. However, the great variability observed in the T cell repertoire which is generated by VDJ recombination and untemplated nucleotide addition to the CDR3 region of TRA and TRB makes it difficult to correlate changes in the repertoire with clinical outcomes. Numerous summary measures such as Shannon’s entropy, Gini index, Euclidean distance and T cell clone tracking strategies have been developed to quantify the differences between donor and recipient T cell profiles, but none have thus far yielded a consistently reliable measure of alloreactivity/immune failure, often because the effects are mediated by polyclonal repertoire shifts occurring in the broader context of numeric T cell population changes. These polyclonal cross repertoire changes are not necessarily fully captured by any of the summary methods currently utilized.

In this paper, a technique widely utilized in signal processing is applied to understand T cell repertoire changes following transplantation, using the donor to recipient transitions in T cell repertoire as a model system in two different patient cohorts. Fourier analysis transforms periodic data in time or spatial domains to the frequency domain, deconstructing complex signals into the constituent frequencies to enable identification of the most important signal components which carry the greatest information or Power in a signal. In essence, transforming the original periodic data into frequencies reveals which wavelength/frequency carries the most information versus trivial details or noise. As an example, in day-to-day life, this helps compress information and in data storage. In evaluating the T cell repertoire, the relative frequencies of the T cell receptor beta (or alpha) V or J segment defined clones may be similarly considered. These TRB and TRA, VDJ recombined sequences, are generated by genomic loci which are logarithmically scaled and periodically distributed. Previous work has demonstrated that spatial ordering the T cell clonal frequencies according to the location of the V or J segments loci on the TRB locus reveals a periodic pattern which is well organized and near identical in pattern in all SCT donors studied, but this ordering is disrupted in SCT recipients. Visually the magnitude of disruption is greater in in patients in different cohorts (ATG vs. PT-CY), who will be at different risk of GVHD, relapse or infections due to varying immune recovery, but no consistent, broad-based objective measure has emerged to help quantify the magnitude of disruption in the T cell repertoire from donor to recipient, across V or J gene segment defined clones, or assess the status of an individual’s immune competence compared with a population normal. Furthermore, the intensity of T cell depletion and immune suppression also restricts the recovering T cell repertoire diversity and consequently the clinical outcomes following transplantation. Thus, it is crucial to understand the magnitude of impact that different immunosuppressive regimens may have on immune recovery in the most objective manner possible.

Spectral (Fourier) analysis of T cell receptor beta V gene defined clonal frequencies examined as a function of the location of the relevant gene segment on the TRB locus (spatial domain) reported herein, demonstrates that in the frequency domain, the normal stem cell donors had a consistent and reproducible spectral pattern of V gene segment usage, for both HLA-matched, related and unrelated individuals. This implies that there is broadly similar use of the TRB V gene segments to yield a diverse, polyclonal repertoire in normal individuals. Furthermore, the relative representation of various V gene segments in the repertoire is dependent on the location of the V (and J) gene segments on the loci. The longer wavelength of spectral centroid and greater value of low frequency indices (LFI) in the donors suggests an organized TCR repertoire, with a hierarchy of V gene segments involved in the recombination process and clonal generation.

An intuitive basis of this notion is such the spectral analysis instead of giving a summary of TCR diversity, by considering relative numbers of individual T cell clones, gives information on the TRB-V gene segment usage across the T cell repertoire. The higher frequency/short wavelength bands indicate more uniform V gene segment usage across the locus and lower frequency/long wavelength bands indicate more scattered and periodic V gene segment usage. Having more of the high frequency bands means a flatter more uniform TRB-V segment distribution amongst the T cell clones, while having an adequate low frequency distribution demonstrates a more normal periodic V gene segment contribution to the repertoire. Spectral centroid tells us the most dominant frequency, and shift in the SC towards a higher frequency means that the repertoire is inclusive of all the V gene segments to a similar extent, which in turn corresponds to a reduction in the low frequency index (and increase in spectral entropy). The opposite situation means V gene segment usage, such that the periodic nature is more evident (lower SC and higher LFI and entropy). Thus, the spectral fingerprint is a way of comparing how close a repertoire is to another, not in terms of its clonal diversity, but in terms of its usage of V gene segments, hence the T cell clones are examined at a lower resolution (exact clonal identity is not required for this perspective). One may think of it as a fingerprint meant to identify and compare repertoires from the point of view of the TRB V gene segment usage, as opposed to the various T cell richness/evenness indices that are calculated, in a sense, it allows distinguishing the forest from the trees. So instead of a measure which gives clonal diversity, this method provides the distinguishing, identifying features of a repertoire based on TRB-V gene segment usage. An analogy may be drawn with emission or absorption spectra in classical physics, where different absorption or emission lines in a normal spectrum allow one to identify elements. Another feature to keep in mind here is the ability of spectral analysis to distinguish shape despite change in magnitude, this is best demonstrated by the PT-CY patients where despite an order of magnitude reduction in the magnitude of T cell clonal frequency, the spatial domain curve and consequently spectral characteristics are maintained and vice versa for the ATG cohort. This is akin to, say a faint Hydrogen emission spectrum from a nearby vs. a distant star, where despite the bright vs. faint spectrum intensity, the spectrum remains unchanged.

Of note, the ATG recipients had a more varied and diffuse power distribution across a broader range of spatial frequencies. The spectral centroid had a higher frequency in general and correspondingly, the LFI was lower. Furthermore, in the ATG recipients the entropy shift was more variable. This is consistent with a more uniform, albeit chaotic and disorderly utilization of TRB V gene segments in the recipient, a less hierarchical VDJ recombination. These polyclonal changes may be attributable to different T cell clones expanding disproportionately and others failing to recover adequately under conditions of alloreactive and pathogen pressure, as well as immune suppression. Eventual T cell reconstitution and in vivo TCR recombination will in such situations be expected to be restorative of the Power spectra to normal. It is possible that patients undergoing other GVHD prophylaxis regimens, such as the non-T cell depleting calcineurin inhibitor and methotrexate may have even greater degrees of perturbation in the ensuing T cell spectra.

Patients who underwent PT-CY administration on the other hand had a different immune recovery trajectory. This was best observed when the power spectra ration between donors and recipients was evaluated. Unlike the ATG recipients, post transplant Power spectra in the recipients following transplant remained essentially unchanged in the PT CY recipients, closely matching the donor spectra. This suggests that the normal pattern of V gene usage. Is restored in the post-transplant setting when CY ablates the alloreactive T cell clones. This selective alloreactive T cell depletion stands in distinction to the indiscriminate T cell depletion wrought by ATG administered pre transplant. These findings, thought provoking as they are, need to be verified in larger cohorts, and should be considered hypothesis generating at this time. Further studies, comparing DRP, which are similar in terms of donor-recipient relatedness, HLA match grade and stem cell source and utilize a similar platform (cDNA or gDNA) for TCR sequencing are necessary for validation. Further, the change vis a vis the background donor spectrum (easily evident in ATG recipients in this analysis) may correspond with different recipient clinical states. Again, larger cohorts of patients are needed to establish such an association.

Given the findings reported herein, one may posit that the donor frequency spectra represent a conserved genomic baseline — a consistent fingerprint of the TRB locus physical structure under conditions of polyclonal normal recombination — while recipient frequency spectra are perturbations of that baseline whose direction and magnitude reflect the clinical state of T-cell reconstitution. Transformation such as the direction of the dominant wavelength change, stable, consistent with preserved repertoire breadth, versus, elongation indicating emergence of oligoclonality will be informative of the state of immune recovery at different times post-transplant. In larger cohorts shifts in spectral characteristics may constitute a spectral fingerprint of immune recovery that is quantifiable and help understand the immune effects of various immune suppressive regimens. Aside from applications in SCT this analytic methodology is likely to enhance the understanding of the T cell effects of solid organ transplant immune tolerance and autoimmune disease.

The major advantage of this analytic methodology is that it provides a way to objectively quantify the robustness of the repertoire recovery, by potentially focusing on the usage of all the TRB gene segments instead of reporting on individual clones, as most clone tracking methods would. It also preserves more information about the T cell repertoire than a single measure such as Shannon’s entropy or Euclidean distance. Utilizing 2-dimensional Fourier Transforms, both the V and J gene segments may be analyzed. The consistency across the frequency spectra in different cohorts of patients, in a manner consistent with prevailing understanding of the mechanism for T cell depletion, supports the dynamical systems nature of T cell responses, which occur in proportion to cognate antigen exposure across the continuum of immune repertoire.

Given the differences in the DRP and T cell sequencing substrate, as well as the small sample size analyzed here with relatively variable time of sample acquisition in the ATG patients, no conclusions may be drawn regarding the clinical utility of this analytic technique. However, the uniformity observed in the donors, and the dispersion seen in the ATG recipients post-transplant, and donor-type recovery seen in PT-CY recipients, it is evident that this methodology will allow a different perspective on immune reconstitution. It is likely that patient with GVHD or CMV reactivation characterized by oligoclonal T cell proliferation will have unique spectral signatures and when measured in the right context this will be a useful tool for discriminating these alloreactive states. Thus, by changing the final pattern of the TRB V gene frequencies, relative expansion of certain clones in the recipient will alter the Power distribution across the constituent frequencies. Similarly, patients with hereditable immune deficiency, autoimmune disease, HIV infections and solid organ transplants may demonstrate variability in the T cell receptor gene segment usage. This may allow discrimination of different clinic states between patients, for example by highlighting oligoclonal autoreactive T cell expansion.

This is an initial proof of concept work and there is much room to improve upon the analytic pipeline presented here, in terms of the best frequency binning for the FFT (recognizing that ideally, it is an infinite continuum), optimization of downstream analytics such as Power spectra and their SC, LFI and band power distribution, spectral entropy, as well as appropriate scaling of the spatial parameters (whether Kilobases or radians), incorporation of the J segment information and so on. A larger cohort with sequencing based on contemporary TRA and TRB clonal definition will be best to accomplish this. Additionally, further work on elucidating the biologic, structural basis of the dominant wavelengths observed in the Power Spectra will be of great interest in terms of the underlying biologic principles governing the evolution of immune responses.

## Data Availability

All data produced in the present study are available upon reasonable request to the authors

## Acknowledgements

The authors gratefully acknowledge Mr. Abdullah A. Toor for his work in collecting the TRB-V gene segment coordinates and Dr. Jeremy Meier for organizing the VCU and John Hopkins University data sets. We also acknowledge Dr. Leo Luznik for a critical review of the manuscript and thoughtful recommendations. Artificial intelligence systems, Claude and ChatGPT were used for the initial Lomb-Scargle spectral calculations and proof of concept development.

## Author attribution

AT: Developed the concept, organized and analyzed the data, wrote the paper; AMV: Analyzed the data and wrote the paper; RQ: Analyzed the data and wrote the paper.

## Supplementary Methods

### Jensen-Shannon Divergence

The Jensen-Shannon Divergence (JSD) was computed between each donor-recipient pair to quantify the overall dissimilarity of their normalized power spectra. JSD is a symmetrized version of the Kullback-Leibler divergence, bounded between 0 (identical spectra) and ln(2) (maximally different). Unlike single-metric comparisons (e.g., SC or band power), JSD captures differences across all frequency bins simultaneously. Both natural and binary (log2) units are reported.

### Bootstrap Confidence Intervals

To quantify uncertainty arising from the limited number of V gene observations (n = 65 or for isolated ATG cohort analysis, 67), non-parametric bootstrap resampling was performed. For each sample, 1000 bootstrap replicates were generated by resampling the V gene observations with replacement. The Lomb-Scargle periodogram and all spectral metrics were recomputed for each replicate. The 95% confidence interval was taken as the 2.5th and 97.5th percentiles of the bootstrap distribution, reported alongside the median.

### Sensitivity Analysis

The sensitivity of the band power metric was assessed by varying two key analytical parameters: the number of frequency bins (128, 512, or 1024) and the minimum frequency cutoff, f_min_ (1/span, 1/120 kb, or 1/60 kb). Six combinations were evaluated for each pair. This analysis establishes the robustness of band-level summary measures to grid resolution and frequency window choice.

### Supplementary Sensitivity Analyses

Two supplementary analyses were performed to verify that specific data features do not artifactually drive the spectral results. First, a pseudogene comb-filter test excluded all 18 TRB V pseudogenes (retaining 49 functional V genes) and recomputed all metrics. This test addresses the concern that zero-frequency data points may introduce a periodic artifact into the power spectrum. Second, a V30 exclusion sensitivity analysis tested whether the inclusion of TRB V30 — which lies 3’ of the TRB D1 segment at position −12.6 kb and was excluded from the original manuscript’s primary analysis — meaningfully alters the spectral metrics.

## Supplementary Results

### Sensitivity Analysis for the ATG cohort

The 3-12 kb band power was computed across three grid sizes (128, 512, and 1024 bins) at two frequency cutoffs (f_min_ = 1/span and f_min_ = 1/60 kb) (Table 4). At a fixed f_min_, increasing the number of bins from 128 to 1024 changed the band power by less than 2 percentage points in all pairs, demonstrating that the band-level metric is essentially invariant to grid resolution once a reasonable minimum is reached.

**Table 4.** Sensitivity Analysis — 3-12 kb Band Power in Donors Across Six Settings.

| Setting | DR3 | DR5 | DR7 | DR8 | DR9 | DR14 |
| --- | --- | --- | --- | --- | --- | --- |
| 128 bins,<br>$f_{\min}=1/\text{span}$ | 48.3% | 42.7% | 52.7% | 43.5% | 48.4% | 44.9% |
| 512 bins,<br>$f_{\min}=1/\text{span}$ | 49.6% | 44.0% | 53.9% | 44.7% | 49.6% | 46.0% |
| 1024 bins,<br>$f_{\min}=1/\text{span}$ | 49.9% | 44.2% | 54.1% | 45.0% | 49.8% | 46.3% |
| 1024 bins,<br>$f_{\min}=1/120$ | 55.2% | 48.8% | 58.1% | 49.4% | 54.4% | 50.4% |
| 1024 bins,<br>$f_{\min}=1/60$ * | 59.7% | 51.8% | 60.4% | 52.6% | 57.9% | 52.9% |
| 128 bins,<br>$f_{\min}=1/60$ | 59.7% | 51.9% | 60.6% | 52.7% | 58.1% | 53.1% |

\* Primary analysis setting. The near-identity of the last two rows (mean delta = 0.15 pp) confirms that the standardized $f_{\min} = 1/60$ kb, rather than grid resolution, is the primary determinant of band power.

Raising f_min_ from 1/span (∼0.0029 cycles/kb, lambda _max_ ∼344 kb (wavelength)) to 1/60 kb (0.0167 cycles/kb, lambda _max_ = 60 kb) increased the 3-12 kb band power by 6-10 percentage points across pairs. This shift is expected: excluding the very low frequencies (wavelengths >60 kb) reallocates that power to the remaining bands, primarily the 3-12 kb and <3 kb bands. The manuscript’s reported 3-12 kb band power of 50-56% is reproduced at the standardized f_min_ = 1/60 kb (51.8-60.4% in our analysis), confirming that the earlier discordance with the manuscript was entirely a function of the frequency window, not a biological difference.

### Jensen-Shannon Divergence

The Jensen-Shannon Divergence, which quantifies the overall dissimilarity of the normalized power spectra between paired donors and recipients, ranged from 0.18 (DR14) to 0.35 (DR3) in natural units (**Table 5**). The ranking of pairs by JSD is: DR3 > DR9 > DR5 ∼ DR7 > DR8 > DR14. This ranking suggests that JSD captures variation in repertoire perturbation even when individual spectral metrics lack statistical significance. The Kullback-Leibler (KL) divergences were roughly symmetric in all pairs (KL(D to R) ∼ KL(R to D)), indicating that the spectral differences are not systematically directional in one condition. This is consistent with the similar band power distributions between donors and recipients.

**Table 5.** Jensen-Shannon Divergence between paired spectra.

| Pair | JSD (natural) | JSD (log2) |
| --- | --- | --- |
| DR3 | 0.3461 | 0.4157 |
| DR5 | 0.2609 | 0.3133 |
| DR7 | 0.2378 | 0.2856 |
| DR8 | 0.2120 | 0.2546 |
| DR9 | 0.2849 | 0.3421 |
| DR14 | 0.1839 | 0.2208 |

*JSD = 0 indicates identical spectra; higher values indicate greater divergence.*

### Pseudogene Comb-Filter Test in the ATG cohort

Eighteen TRB V genes in this dataset are pseudogenes (marked with asterisks), having clonal frequency = 0 but occupying biologically relevant genomic positions. To test whether these zero-frequency data points introduce a periodic artifact (comb filter) into the power spectrum, the full pipeline was re-run after excluding all 18 pseudogenes, retaining 49 functional V genes.

Excluding pseudogenes changed the 3-12 kb band power by +0.6 to +1.1 percentage points across pairs (**Supplementary Table A**) and the spectral centroid by less than 0.7 millicycles/kb in all cases. The spectral fingerprint is therefore not driven by pseudogene artifacts and reflects genuine V-gene usage patterns in functional genes.

**Supplementary Table A.** Pseudogene Exclusion — Effect on 3-12 kb Band Power.

| Pair | All 67 genes (%) | No pseudogenes (49 genes, %) | Delta (pp) |
| --- | --- | --- | --- |
| DR3 | 59.7 | 60.3 | +0.6 |
| DR5 | 51.8 | 52.8 | +1.0 |
| DR7 | 60.4 | 61.3 | +1.0 |
| DR8 | 52.6 | 53.7 | +1.1 |
| DR9 | 57.9 | 58.6 | +0.7 |
| DR14 | 52.9 | 53.9 | +1.0 |

### V30 Inclusion/Exclusion Sensitivity test for the ATG cohort

Our primary analysis includes V30 for completeness. To verify that this does not affect the results, the pipeline was re-run excluding V30 (retaining 66 V genes). Excluding V30 changed the 3-12 kb band power by −0.2 to −0.9 percentage points, and the SC by −1.5 to −4.3 millicycles/kband JSD by −0.010 to +0.011 (**Supplementary Table B**). None of these changes are analytically meaningful, and the JSD ranking of pairs (DR3 > DR9 > DR5 > DR7 > DR8 > DR14) is preserved.

**Supplementary Table B.**
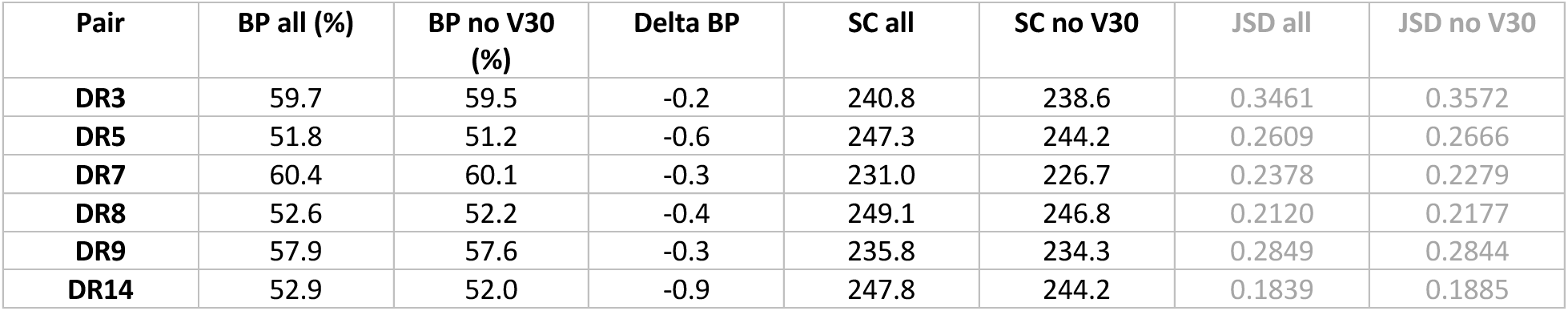
V30 Exclusion — Effect on Key Spectral Metrics.

| Pair | BP all (%) | BP no V30 (%) | Delta BP | SC all | SC no V30 | JSD all | JSD no V30 |
| --- | --- | --- | --- | --- | --- | --- | --- |
| **DR3** | 59.7 | 59.5 | -0.2 | 240.8 | 238.6 | 0.3461 | 0.3572 |
| **DR5** | 51.8 | 51.2 | -0.6 | 247.3 | 244.2 | 0.2609 | 0.2666 |
| **DR7** | 60.4 | 60.1 | -0.3 | 231.0 | 226.7 | 0.2378 | 0.2279 |
| **DR8** | 52.6 | 52.2 | -0.4 | 249.1 | 246.8 | 0.2120 | 0.2177 |
| **DR9** | 57.9 | 57.6 | -0.3 | 235.8 | 234.3 | 0.2849 | 0.2844 |
| **DR14** | 52.9 | 52.0 | -0.9 | 247.8 | 244.2 | 0.1839 | 0.1885 |

**Supplementary Figure P1:**
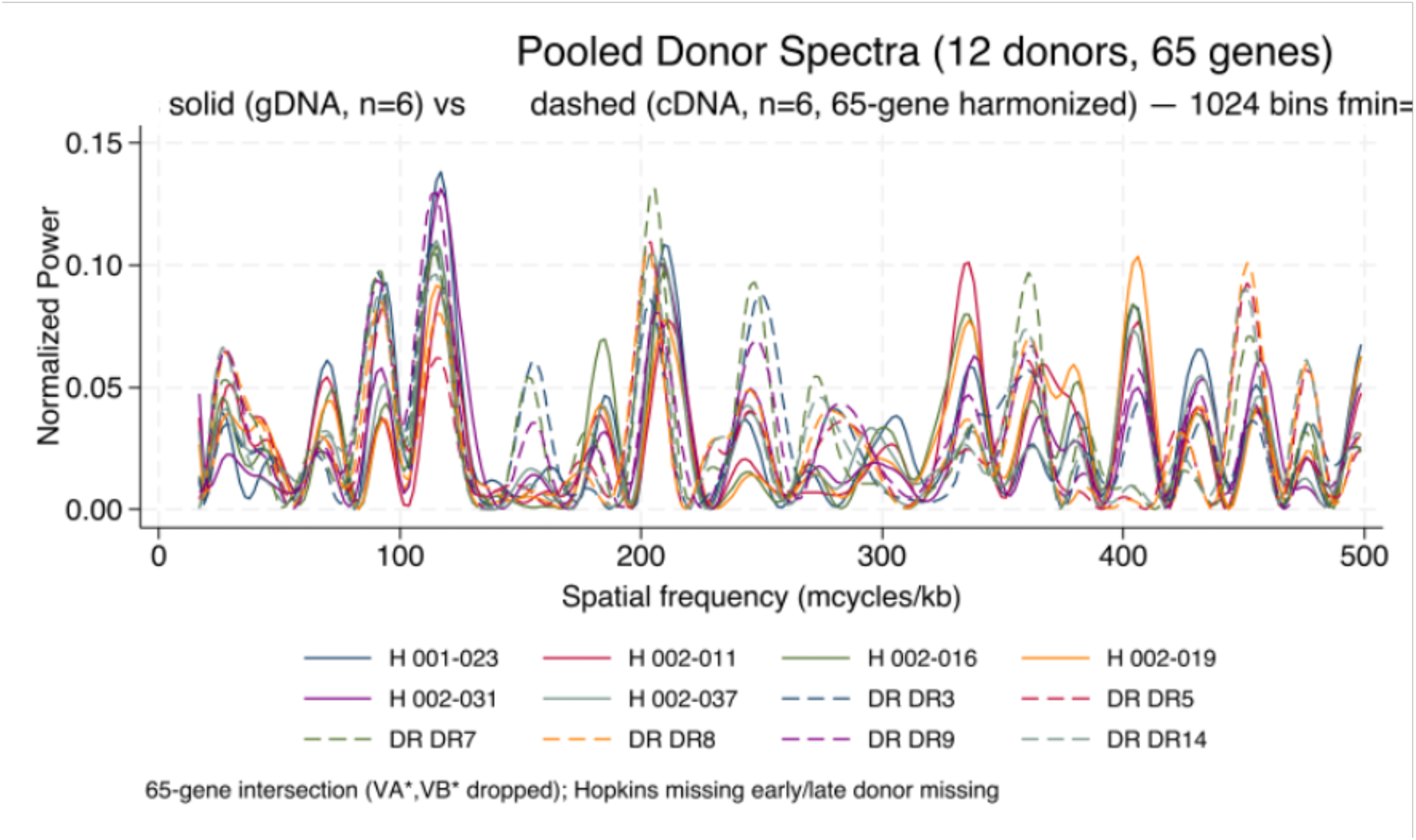
Pooled donor spectra (12 donors, 65 genes, solid PT-CY cohort gDNA vs dashed ATG cohort cDNA). All 12 share dominant 3–12 kb peaks 115–335 mcycles/kb; Hopkins donors slightly higher power at 200–300 mcycles/kb, DR donors slightly higher at <80 mcycles/kb.

**Supplementary Figure P2:**
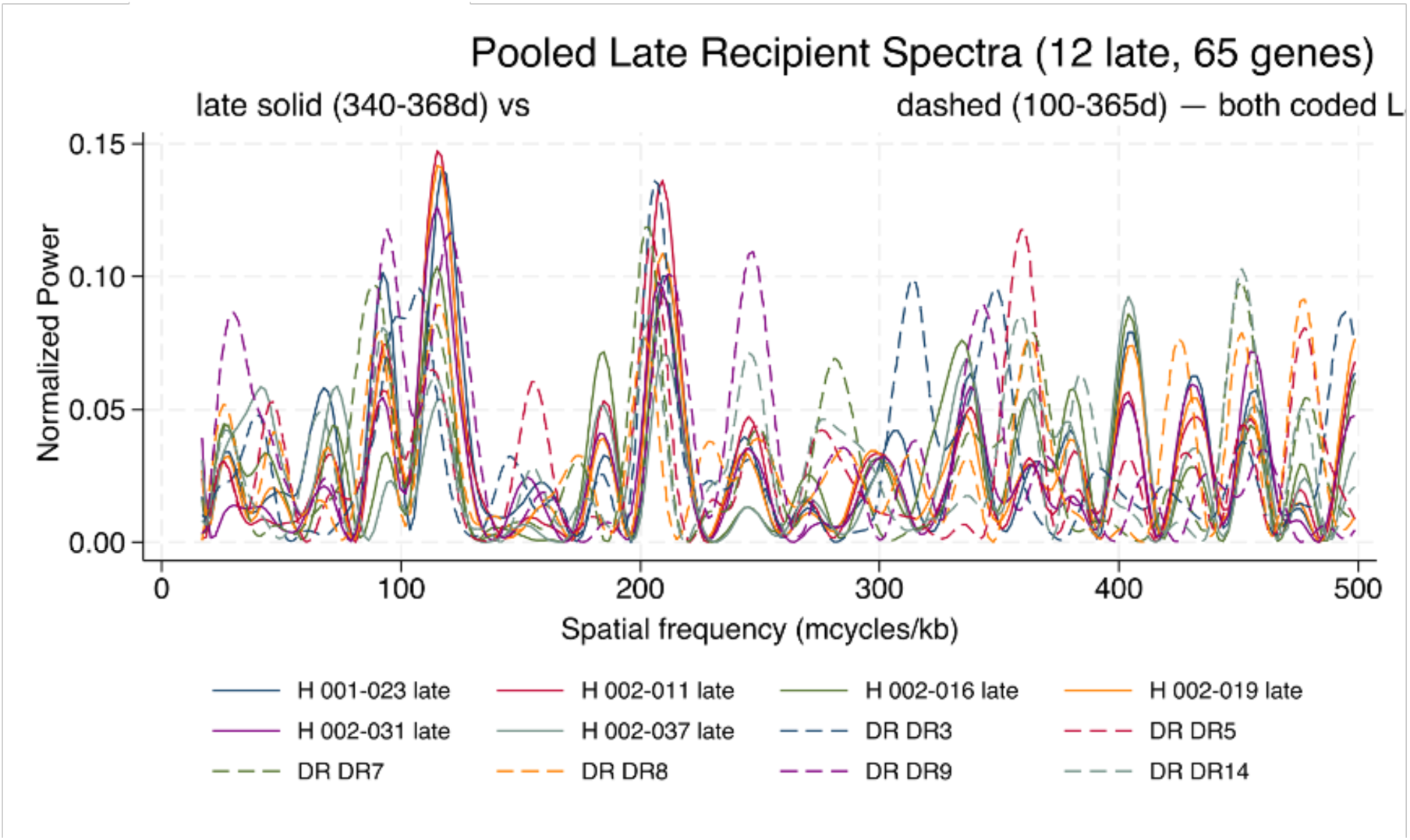
Pooled late recipient spectra (12: 6 Hopkins late +6 DR R). Greater spread than donors, especially DR late (dashed) more variable above 100 mcycles/kb, echoing isolated late divergence.

**Supplementary Figure P3:**
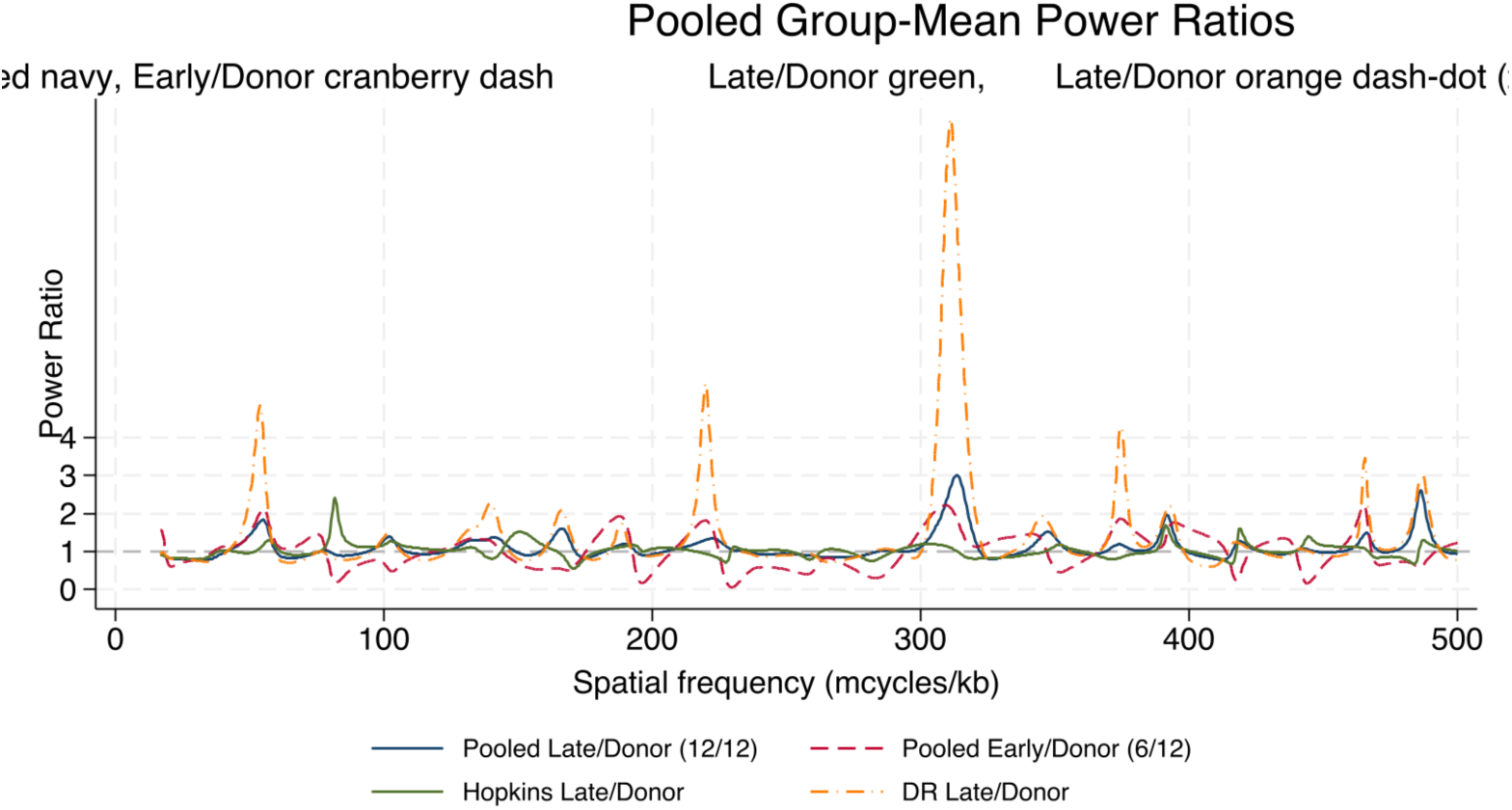
Pooled group-mean power ratios. PT CY Late/Donor pooled navy slightly >1 at 100– 200 mcycles/kb (6–10 kb) and <1 at <30 mcycles/kb; PT CY Early/Donor cranberry dash similar; PT CY Late/Donor green vs ATG Late/Donor orange dash-dot diverge at >300 mcycles/kb.

## References

1 Meier J, Roberts C, Avent K, Hazlett A, Berrie J, Payne K, Hamm D, Desmarais C, Sanders C, Hogan KT, Archer KJ, Manjili MH, Toor AA. Fractal organization of the human T cell repertoire in health and after stem cell transplantation. Biol Blood Marrow Transplant. 2013 Mar;19(3):366–77. doi: 10.1016/j.bbmt.2012.12.004. Epub 2013 Jan 11. PMID: 23313705.

2 https://www.ncbi.nlm.nih.gov/gene/6957

3 Toor AA, Toor AA, Rahmani M, Manjili MH. On the organization of human T-cell receptor loci: log-periodic distribution of T-cell receptor gene segments. J R Soc Interface. 2016 Jan;13(114):20150911. doi: 10.1098/rsif.2015.0911. PMID: 26763333; PMCID: PMC4759796.

4 Meier JA, Haque M, Fawaz M, Abdeen H, Coffey D, Towlerton A, Abdeen A, Toor A, Warren E, Reed J, Kanakry CG, Keating A, Luznik L, Toor AA. T Cell Repertoire Evolution after Allogeneic Bone Marrow Transplantation: An Organizational Perspective. Biol Blood Marrow Transplant. 2019 May;25(5):868–882. doi: 10.1016/j.bbmt.2019.01.021. Epub 2019 Jan 21. PMID: 30677510; PMCID: PMC6645918.

5 Abdul Razzaq B, Scalora A, Koparde VN, Meier J, Mahmood M, Salman S, Jameson-Lee M, Serrano MG, Sheth N, Voelkner M, Kobulnicky DJ, Roberts CH, Ferreira-Gonzalez A, Manjili MH, Buck GA, Neale MC, Toor AA. Dynamical System Modeling to Simulate Donor T Cell Response to Whole Exome Sequencing-Derived Recipient Peptides Demonstrates Different Alloreactivity Potential in HLA-Matched and -Mismatched Donor-Recipient Pairs. Biol Blood Marrow Transplant. 2016 May;22(5):850–61. doi: 10.1016/j.bbmt.2015.11.1103. Epub 2015 Dec 11. PMID: 26688192; PMCID: PMC4955725.

6 Koparde V, Abdul Razzaq B, Suntum T, Sabo R, Scalora A, Serrano M, Jameson-Lee M, Hall C, Kobulnicky D, Sheth N, Feltz J, Contaifer D Jr, Wijesinghe D, Reed J, Roberts C, Qayyum R, Buck G, Neale M, Toor A. Dynamical system modeling to simulate donor T cell response to whole exome sequencing-derived recipient peptides: Understanding randomness in alloreactivity incidence following stem cell transplantation. PLoS One. 2017 Dec 1;12(12):e0187771. doi: 10.1371/journal.pone.0187771. PMID: 29194460; PMCID: PMC5711034.

7 Toor AA, Sabo RT, Roberts CH, Moore BL, Salman SR, Scalora AF, Aziz MT, Shubar Ali AS, Hall CE, Meier J, Thorn RM, Wang E, Song S, Miller K, Rizzo K, Clark WB, McCarty JM, Chung HM, Manjili MH, Neale MC. Dynamical System Modeling of Immune Reconstitution after Allogeneic Stem Cell Transplantation Identifies Patients at Risk for Adverse Outcomes. Biol Blood Marrow Transplant. 2015 Jul;21(7):1237–45. doi: 10.1016/j.bbmt.2015.03.011. Epub 2015 Apr 4. PMID: 25849208; PMCID: PMC4836381.

8 Zelikson V, Sabo R, Serrano M, Aqeel Y, Ward S, Al Juhaishi T, Aziz M, Krieger E, Simmons G, Roberts C, Reed J, Buck G, Toor A. Allogeneic haematopoietic cell transplants as dynamical systems: influence of early-term immune milieu on long-term T-cell recovery. Clin Transl Immunology. 2023 Jul 13;12(7):e1458. doi: 10.1002/cti2.1458. PMID: 37457614; PMCID: PMC10345185.

9 Meier J, Roberts C, Avent K, Hazlett A, Berrie J, Payne K, Hamm D, Desmarais C, Sanders C, Hogan KT, Archer KJ, Manjili MH, Toor AA. Fractal organization of the human T cell repertoire in health and after stem cell transplantation. Biol Blood Marrow Transplant. 2013 Mar;19(3):366–77. doi: 10.1016/j.bbmt.2012.12.004. Epub 2013 Jan 11. PMID: 23313705.

10 Kanakry CG, Coffey DG, Towlerton AM, Vulic A, Storer BE, Chou J, Yeung CC, Gocke CD, Robins HS, O’Donnell PV, Luznik L, Warren EH. Origin and evolution of the T cell repertoire after posttransplantation cyclophosphamide. JCI Insight. 2016;1(5):e86252. doi: 10.1172/jci.insight.86252. Epub 2016 Apr 21. PMID: 27213183; PMCID: PMC4874509.

11 Meier JA, Haque M, Fawaz M, Abdeen H, Coffey D, Towlerton A, Abdeen A, Toor A, Warren E, Reed J, Kanakry CG, Keating A, Luznik L, Toor AA. T Cell Repertoire Evolution after Allogeneic Bone Marrow Transplantation: An Organizational Perspective. Biol Blood Marrow Transplant. 2019 May;25(5):868–882. doi: 10.1016/j.bbmt.2019.01.021. Epub 2019 Jan 21. PMID: 30677510; PMCID: PMC6645918.

12 Sampson, J.K., Sheth, N.U., Koparde, V.N., Scalora, A.F., Serrano, M.G., Lee, V., Roberts, C.H., Jameson-Lee, M., Ferreira-Gonzalez, A., Manjili, M.H., Buck, G.A., Neale, M.C. and Toor, A.A. (2014), Whole exome sequencing to estimate alloreactivity potential between donors and recipients in stem cell transplantation. Br J Haematol, 166: 566–570. 10.1111/bjh.12898

